# Characterization of the Subclinical Perilesional Zone in the Macula of Early-Stage ABCA4 Disease

**DOI:** 10.1101/2024.11.16.24317331

**Authors:** Aiden Zee, Winston Lee, Pei-Yin Su, Jana Zernant, Stephen H. Tsang, Rando Allikmets

## Abstract

**Purpose:** To characterize photoreceptor layer thinning in clinically unremarkable regions adjacent to the atrophic lesion in early-stage ABCA4 disease eyes.

**Methods:** 27 patients with confined atrophic lesions (*≤*3.5mm in diameter) were included. Two pathogenic alleles were confirmed by sequencing of the *ABCA4* locus. Multimodal imaging included color fundus photography, short wavelength-autofluorescence (SW-AF) and near infrared-autofluorescence (NIR-AF) imaging. Total receptor+ (TREC+) thickness was segmented in spectral domain-optical coherence tomography (SD-OCT) scans in patient eyes (n=27) along with age-matched healthy control eyes (n=20).

**Results:** *µ*_*age*_ of the study cohort was 24.1 years and 15/27 (55.6%) patients harbored genotypes consisting of the p.(Gly1961Glu) variant in *ABCA4*. Atrophic lesions in the central macula ranged from 0.61 to 3.13 mm in diameter (*µ* = 1.73, *σ* = 0.70). Six patients had mild RPE mottling adjacent to the lesion on NIR-AF. The atrophic lesion corresponded to a disruption of photoreceptor-attributable bands on SD-OCT while all layers were visibly intact outside the lesion. TREC+ thickness in patient eyes were <0.15 mm or below 4*σ* of normal control eyes immediately adjacent to the lesion edge and gradually normalized to within *±* 2*σ* at *≈* 1.2 mm eccentricity from the fovea.

**Conclusion:** A uniform subclinical perilesional zone (SPZ) of photoreceptor thinning extends around the perimeter of early-stage atrophic lesions in ABCA4 disease. This region spatially maps to known regions of vision loss and more accurately approximates the extent of photoreceptor abnormality compared to the disease changes visible on standard fundus imaging.

**Translational relevance:** Semi-automated segmentation of SD-OCT scans identifies a consistent subclinical biomarker relevant to early photoreceptor degeneration in ABCA4 disease.

## 1 Introduction

Pathogenic variation in the *ABCA4* gene (MIM*♯* 601691) is the underlying cause[1] of several retinal degenerative disorders including Stargardt disease,[74] fundus flavimaculatus,[16, 21, 23, 24] cone-rod dystrophy (CRD),[22, 49] rapid-onset chorioretinopathy (ROC),[83] late-onset maculopathy,[42, 62, 61, 95, 94] among many others.[13] Each of these disorders is characterized by the progressive degeneration of photoreceptors, specialized light-sensitive cells in the retina responsible for vision. The ABCA4 protein plays a crucial role in the visual cycle by transporting byproducts, specifically N-retinylidene-PE,[3] out of photoreceptors and into retinal pigment epithelium (RPE) cells. Impaired ABCA4 function impedes this clearance process resulting in the irreversible formation and subsequent accumulation[6, 11] of cytotoxic bisretinoids (lipofuscin) that trigger the death of both the RPE photoreceptor layers in the retina.[73, 69, 70]

Clinically, this degenerative process manifests as an atrophic lesion in the central macula that expands outwardly from the fovea over time resulting in further deterioration of central vision.[13] While the size of the atrophic lesion in the retina generally correlates with the severity of vision loss, prior studies have found that the outer boundary of the dense scotoma (blind spot) typically extends beyond the visible edge of the atrophic lesion in the fundus—a finding that is unique to ABCA4 disease as compared to other macular disorders such as dry age-related macular degeneration (AMD).[56, 81] Consistent with this phenomenon, fixation studies have shown that the eccentric preferred retinal locus (PRL) in ABCA4 disease eyes is located far away from the edge of the atrophic lesion.[27, 59, 64, 65, 86] Furthermore, clinically unremarkable regions adjacent to the atrophic lesion have also been shown to exhibit reduced visual sensitivity by psychophysical tests such as fundus-driven perimetry [4, 10, 80, 79] and decreased electrophysiological function on multifocal electroretinogram (mfERG) recordings.[39, 40, 66]

Although early functional attenuation in otherwise “clinically unremarkable” areas of the fundus has been observed in ABCA4 disease,[2, 85] more sophisticated imaging techniques have nevertheless revealed that these areas in fact do exhibit disease-related structural changes. Optical coherence tomography (OCT) is a non-invasive imaging technique that captures high-resolution, cross-sectional images of the retina, allowing for detailed visualization of its layers and detection of subtle structural changes in eye diseases.[29] Lesions marked by outer retinal atrophy in the fundus correspond to the loss or disruption of photoreceptor- and RPE-attributable layers visible on individual B-scans. While these layers are generally intact in clinically unremarkable areas, several studies have shown that they exhibit significant thinning compared to healthy eyes.[25, 57, 38, 41, 50, 52, 57, 67, 78, 88] Retinal layer thinning is a commonly recognized precursor stage to atrophy in both patients and animal models of ABCA4 disease,[28, 47, 56, 72] making it a valuable biomarker for tracking disease progression and evaluating clinical trial outcomes. Characterizing retinal layer thinning in ABCA4 disease in a consistent manner, however, has been challenging due to its profound clinical heterogeneity.[13] To accommodate this heterogeneity, most prior studies have focused on broad averaging approaches (comparing fixed regions and ETDRS grids)[25, 26, 38, 50, 52, 67, 78, 88, 89] that obscure localized variations which may be insightful to disease progression. To make better use of retinal layer thinning as a reliable biomarker in the clinic and clinical trials, a more incisive approach focusing on localized and consistent changes relative to specific disease patterns is necessary.

The aim of this study is to characterize localized changes in photoreceptor layer thickness in areas adjacent to atrophic lesions in ABCA4 disease. To minimize the effect of variable disease features that may confound thickness measurements (e.g., the presence of yellow, pisciform flecks[14, 71], QDAF,[75] etc.) study inclusion was limited to early-stage patients/eyes with confined atrophic lesions and no other visible surrounding (extra-lesional) disease alterations in the fundus.

## 2 Materials & Methods

### 2.1 Ethics approval

All study subjects were consented before participation under the protocol #AAAI9906 approved by the Institutional Review Board at Columbia University Irving Medical Center. All study-related procedures adhered to the tenets established by the Declaration of Helsinki.

### 2.2 Clinical examination

Complete ophthalmic examinations were provided by a retinal specialist, which included a slit-lamp and dilated fundus examination. Color fundus photographs (30° and 50° magnification) were captured using the FF450plus Fundus Camera (Carl Zeiss Meditec). Fundus autofluorescence images (both short wavelength-autofluorescence (SW-AF, 488-nm excitation) and near infrared-autofluorescence (NIR-AF, 787-nm excitation)), and high-resolution spectral-domain optical coherence tomography (SD-OCT) scans were acquired using a Spectralis HRA+OCT (Heidelberg Engineering, Heidelberg, Germany).

### 2.3 Molecular screening

All causal genetic variants were identified by either direct sequencing of the *ABCA4* locus or whole-exome sequencing as previously described.[92, 93] All detected possibly disease-associated variants were confirmed by Sanger sequencing and determined to be pathogenic through multiples lines of evidence (Supplemental Table S1). Variants identified from sequencing were annotated with established pathogenicity prediction scores, including phyloP100,[58] M-CAP (percentile range 0 to 1),[32] REVEL (likelihood ratio 0 to 1),[30] CADDv1.6 (PHRED scale 0 to 48),[37, 60] DANN, Eigen (probability score -4.09 to 6.31),[31] SpliceAI (Δscore)[33] and Pangolin (Δscore),[91] using ANNOVAR.[87] Minor allele frequences (MAF) were obtained from the gnomAD database v3.1.2 (https://gnomad.broadinstitute.org/gene/ENSG00000198691?dataset=gnomad_r4) (assessed August 2024). Finally, the phase of all variants were either confirmed by parental screening or imputed based on co-occurrence in the general populations (https://gnomad.broadinstitute.org/variant-cooccurrence).

### 2.4 Outer retinal layer segmentation on SD-OCT

Retinal layer thicknesses were obtained by segmenting single 9-mm high resolution SD-OCT scans through the fovea in a single, randomly selected eye from each patient and 20 age-matched, healthy control eyes (Supplemental Figure S1). Total receptor+ (TREC+) was defined as the distance between the Bruch’s membrane/choroid interface and the inner nuclear layer (INL)/outer plexiform layer (OPL) boundary (Supplemental Figure S2).[28] The boundaries of TREC+ were measured manually using the caliper tool in the HEYEX software (Heidelberg Engineering, Heidelberg, Germany). The thickness of TREC+ was measured at 0.5 mm interval positions (13 positions total) along the temporal and nasal meridian from the fovea (origin). In plots, all thickness profiles are represented as right eyes.

### 2.5 Statistical analyses

Statistical analyses and plots (mean of grader measurements) were performed using a custom written program in R (https://www.r-project.org) through RStudio (https://www.rstudio.com). SD-OCT measurements were performed by two independent graders (AZ and WL) along with a third adjudicator when necessary (PYS). Intraclass correlation coefficients (ICC), calculated using the irrICC package (https://cran.r-project.org), showed excellent intergrader agreement for patients (ICC>0.9818) and controls (ICC>0.9997). Comparison of non-normal distributions (e.g. age at examinaation, fundus measurements) were based on unpaired, two-tailed P-values from Mann-Whitney U test. Wilcoxon signed rank test for paired data was used oo test the symmetry of values from the same eye across patients (e.g., temporal and nasal SPZ width). Contingency tables (2×2) were compared using both a Fisher’s exact test and Chi-square test with Yate’s correction.

## 3 Results

### 3.1 Clinical examination and genetic screening

Twenty-seven patients with confined atrophic lesions and no other surrounding disease alterations in the macula detected on fundus examination were included in the study (Figure 1). Considering the imaging field of the testing modalities (see Methods), patients/eyes with lesions with diameters of >3.5 mm (area *≈* 10 *mm*^2^) were excluded to ensure that an adequate area of the adjacent fundus was available for analysis. Demographic, clinical and genetic characteristics of the study cohort are summarized in Table 1. Cohort age ranged from 11-65 years; mean (*µ*) = 24.1 years, standard deviation (*σ*) = 12.0 years.

**Table 1:**
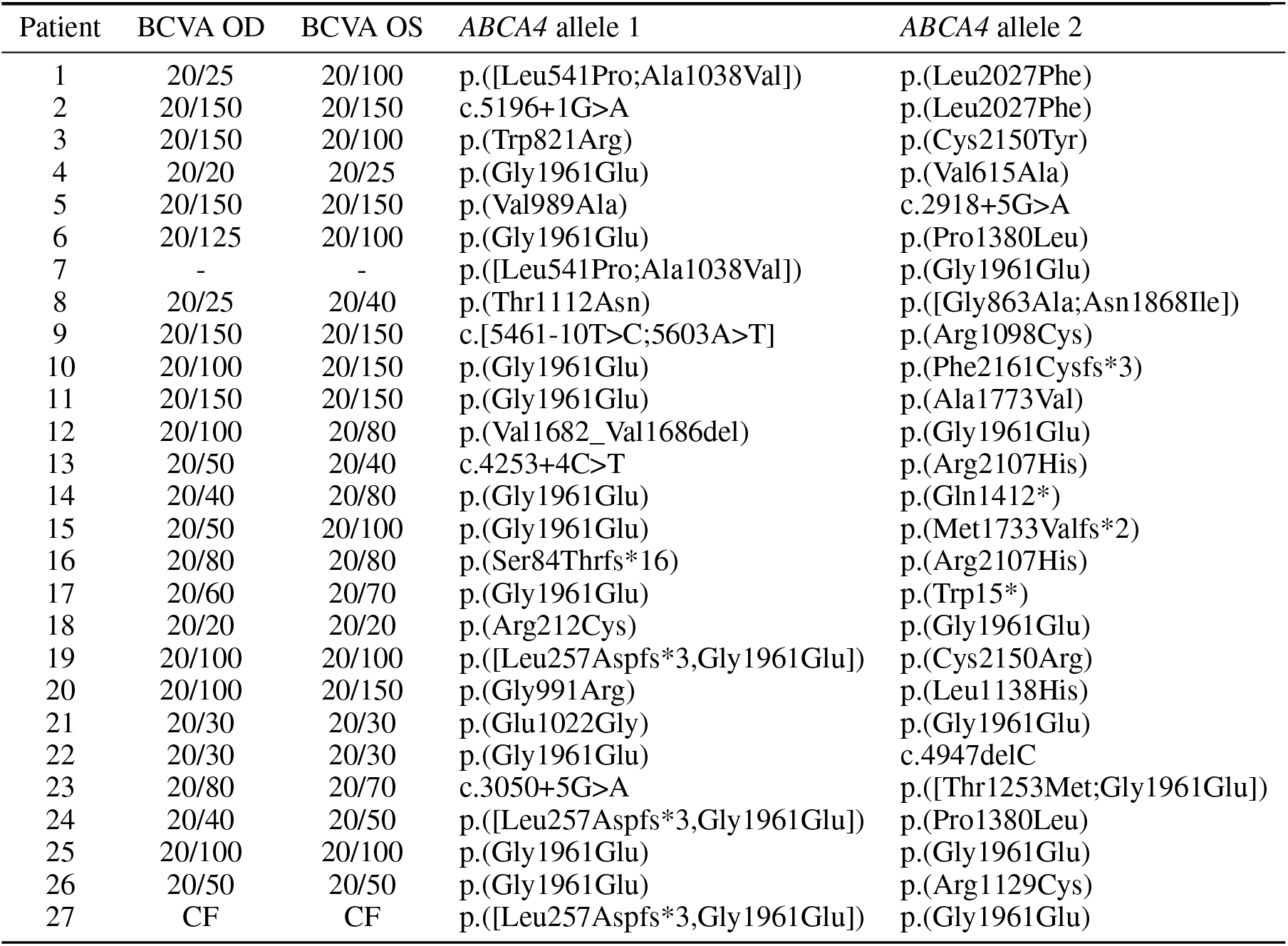
Demographic, clinical and genetic characteristics of the study cohort.

**Figure 1.**
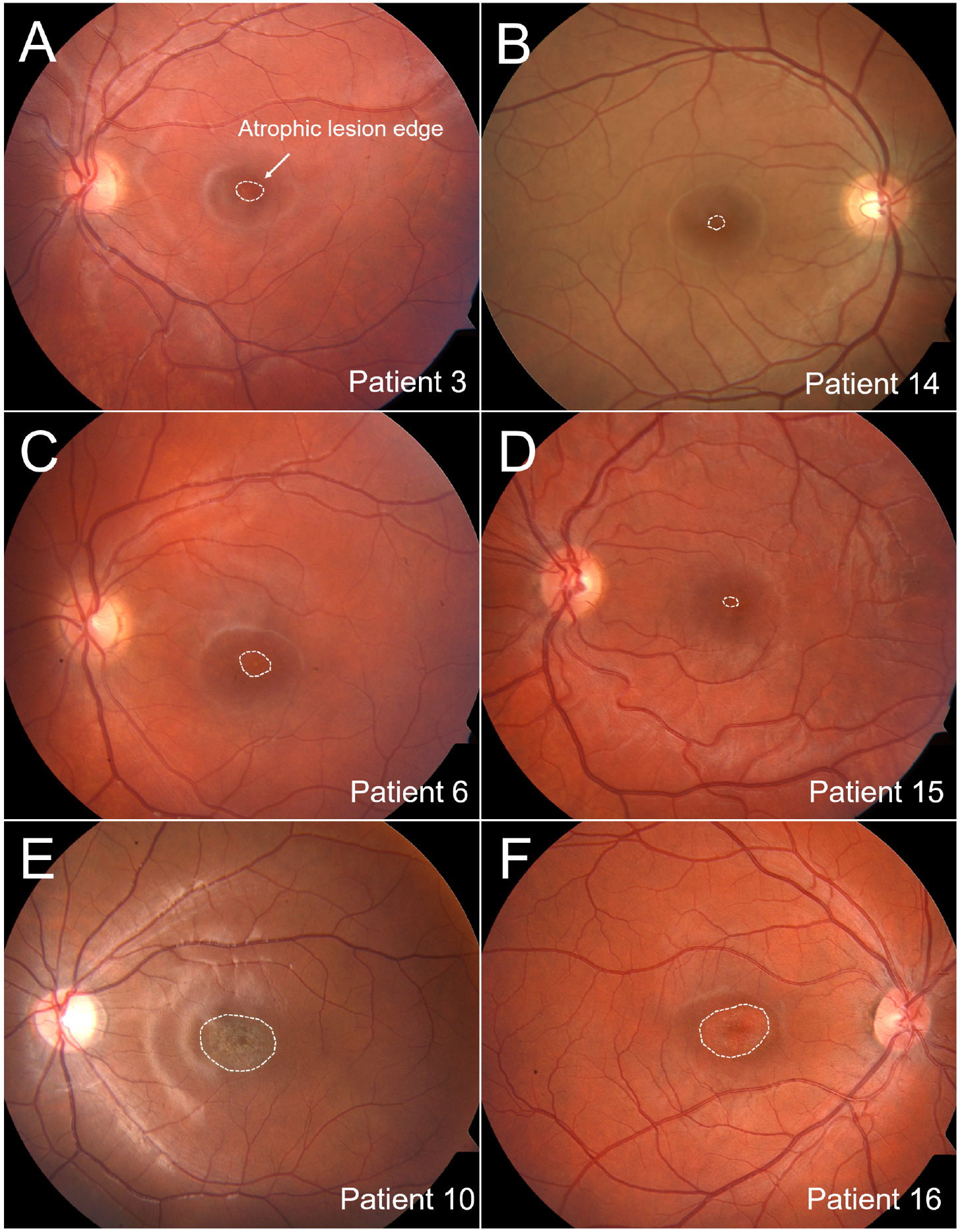
Color fundus photographs showing the confined atrophic lesion and clinically unremarkable periphery of the (A)left eye of Patient 3, (B) right eye of Patient 14, (C) left eye of Patient 6, (D) left eye of Patient 15, (E) left eye of Patient 10 and (F) right eye of Patient 16. The white dotted line delineates the visible edge of each atrophic lesion.

### 3.2 Sequencing and analysis of *ABCA4*

All patients were biallelic for known or expected disease-causing variants in *ABCA4*. Phase was confirmed by familial segregation in all except 4 patients (P16, P23, P24 and P26); phasing in the latter was statistically imputed based on co-occurrence in gnomAD (Supplemental Figure S3). Thirty-three unique variants were identified across the cohort: 17 missense, 5 complex alleles, 2 nonsense, 5 intronic substitutions, 3 deletions and 1 duplication. Predicted in silico or established pathogenicity evidence for each unique variant are summarized in Supplemental S1. The phenotype selected in this study is consistent with the clinical diagnosis of bull’s eye maculopathy (BEM), an entity associated with the frequent p.(Gly1961Glu) mutation.[8] As expected, therefore, the majority of genotypes in this study cohort (15/27 patients, 56%) consisted of the p.(Gly1961Glu) variant. Notably, four of the p.(Gly1961Glu) alleles harbored additional variants in cis: three with the deep intronic c.769-784C>T modifier[43, 61, 63] (P19, P24 and P27) and one with the p.(Thr1253Met) coding variant (P23).

### 3.3 Multimodal fundus autofluorescence (FAF) imaging

The atrophic lesion observed on fundoscopy appeared as a dark region on both fundus autofluorescence modalities (Figure 2). On short wavelength-autofluorescence (SW-AF, 488-nm excitation), the central hypoAF region exhibited a heterogeneous (punctate) appearance in most eyes (38/54 eyes, 70%) and was surrounded by a hyperAF border in 28 eyes (52%). Comparatively, the central hypoAF lesion exhibited a more homogeneous appearance on near infrared -autofluorescence (NIR-AF, 787-nm excitation). On both modalities, no significant disease-associated abnormalities were detected beyond the lesion boundary except in a subgroup of 6 patients (P1, P8, P13, P19, P23 and P24) in whom mild RPE mottling immediately adjacent to the lesion border was visible on NIR-AF (Figure 2C and Figure 22D).

**Figure 2.**
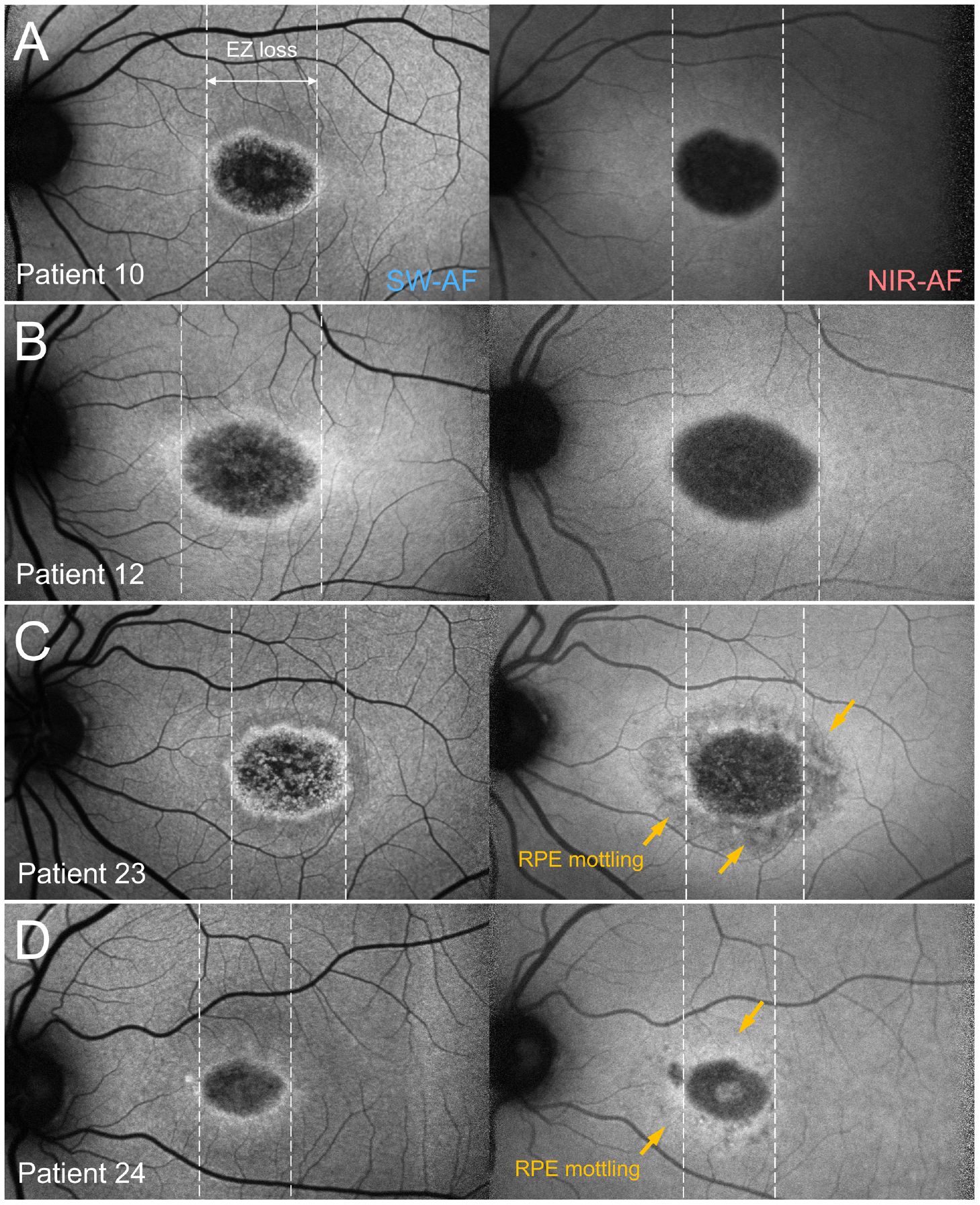
Short wavelength-autofluorescence (SW-AF) (left column) and near infrared-autofluorescence (NIR-AF) (right column) imaging of the atrophic lesion in the left eyes of (A) Patient 10, (B) Patient 12, (C) Patient 23 and (D) Patient 24. White, horizontal dotted lines mark the position of ellipsoid zone (EZ) loss within the atrophic lesion. Yellow arrows indicate perilesional RPE mottling on NIR-AF.

### 3.4 Spectral-domain optical coherence tomography (SD-OCT)

Within the atrophic lesion, several outer retinal layers were visibly disrupted—specifically, the outer retinal layer (ONL), external limiting membrane (ELM) and the ellipsoid zone (EZ) band on SD-OCT (Figure 3). Comparative analysis between FAF and SD-OCT confirmed that the horizontal diameter of the atrophic lesions on both SW-AF and NIR-AF images (Figure 2, dashed vertical lines) spatially corresponds best with the horizontal diameter of EZ loss.[17, 35, 53, 55] The mean horizontal diameter of EZ loss in all study eyes was 1.76 mm (*σ* = 0.70, 0.61-3.13 mm) (Table 2). Although all outer retinal layers adjacent to the atrophic lesion were present and intact, several visible abnormalities were noted in this region. In all study eyes, the ELM was visibly thickened[36, 44] and the interdigitation zone (IZ)[34] was visibly indiscernible between the EZ and RPE layers (Figure 3, insets). In both eyes of 20/27 patients (74%), hyper-reflective foci were observed between the ELM and outer plexiform layer (OPL).[9] Most notably, this entire region appeared to be abnormally thin, particularly the ONL and other photoreceptor-attributable layers, relative to healthy eyes (Figure 3).

**Table 2:** Demographic, clinical and genetic characteristics of the study cohort.

| Patient | Age | Lesion radius |  | Lesion diameter | SPZ width |  | Mean SPZ width |
| --- | --- | --- | --- | --- | --- | --- | --- |
|  |  | Temporal | Nasal |  | Temporal | Nasal |  |
| All study eyes |  |  |  |  |  |  |  |
| Mean | 24.1 | 0.907 | 0.927 | 1.726 | 1.175 | 1.25 | 1.212 |
| Median | 23.9 | 0.995 | 0.98 | 1.881 | 1.032 | 1.167 | 1.157 |
| SD | 12 | 0.351 | 0.333 | 0.701 | 0.476 | 0.358 | 0.379 |
| Other *ABCA4* |  |  |  |  |  |  |  |
| Mean | 20.8 | 0.967 | 0.988 | 1.711 | 1.098 | 1.2 | 1.149 |
| Median | 19.6 | 1 | 0.984 | 1.825 | 0.878 | 1.164 | 1.158 |
| SD | 8.2 | 0.309 | 0.263 | 0.661 | 0.415 | 0.307 | 0.304 |
| p.(Gly1961Glu) |  |  |  |  |  |  |  |
| Mean | 26.7 | 0.86 | 0.878 | 1.738 | 1.237 | 1.29 | 1.263 |
| Median | 24.2 | 0.957 | 0.936 | 1.926 | 1.103 | 1.38 | 1.157 |
| SD | 14.1 | 0.386 | 0.381 | 0.754 | 0.525 | 0.401 | 0.434 |
| Mann-Whitney U test |  |  |  |  |  |  |  |
| P-value | 0.283 | 0.714 | 0.648 | 0.714 | 0.456 | 0.548 | 0.486 |
| Test statistic W | 112 | 82 | 80 | 82 | 106 | 103 | 105 |

**Figure 3.**
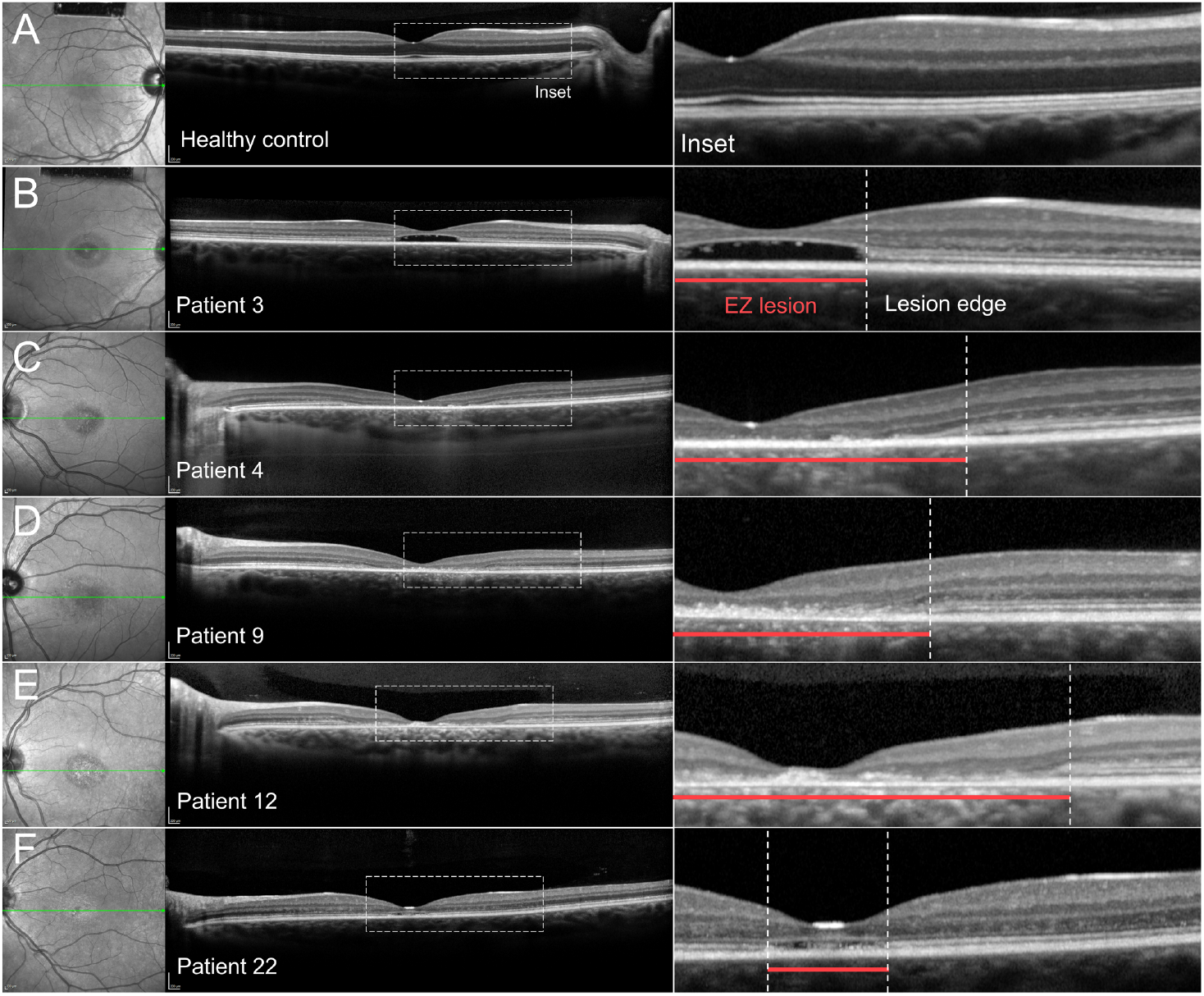
Horizontal spectral domain-optical coherence tomography (SD-OCT, 9mm) scans through the fovea in (A) a healthy control eye (N1), (B) Patient 3, (C) Patient 4, (D) Patient 9, (E) Patient 12 and (F) Patient 22. The corresponding position of each scan is indicated (green line) on infrared reflectance images of the macula. Red horizontal lines delineate the continuous area of EZ lesion and white vertical dotted lines mark the position of EZ disruption.

### 3.5 Photoreceptor layer thickness quantification

To quantify the extent of photoreceptor-attributable layer thinning, the thickness of total receptor+ (TREC+), defined as the distance between the Bruch’s membrane(BM)/choroid(Ch) interface and the inner nuclear layer (INL)/OPL boundary[28] (Supplemental Figure S2), was segmented in all patients (27 eyes) and compared to 20 age-matched, healthy control eyes (Supplemental Table S1) (Supplemental Figure S1). On average, TREC+ thickness fell below -2*σ* approximately 1.5 - 2.0 mm on both the temporal nasal sides of the EZ lesion (Figure 4A) (Table 2). Significant thinning was most pronounced in immediately adjacent areas and gradually normalized (to within ±2*σ* of healthy eyes) with increasing eccentricity. TREC+ thickness in all study eyes were below 4*σ* of healthy eyes at *≈* 0.5 mm from the EZ lesion (Figure 4B). At 1.0 mm eccentricity, significant thinning was still found in 13/27 (48%) study eyes, and at 2.0 mm eccentricity, thickness in all except one (P18) returned to within *±* 2*σ* of healthy eyes (Figure 4B). This region of significant TREC+ thinning (below 2*σ*), referred to henceforth as the subclinical perilesional zone (SPZ), covers a mean distance of 1.18 mm (*σ* = 0.48) and 1.25 mm (*σ* = 0.36) on the temporal and nasal sides of the lesion border, respectively. There was no significant difference between the temporal and nasal SPZ widths (paired Wilcoxon signed rank test, P = 0.215964).

**Figure 4.**
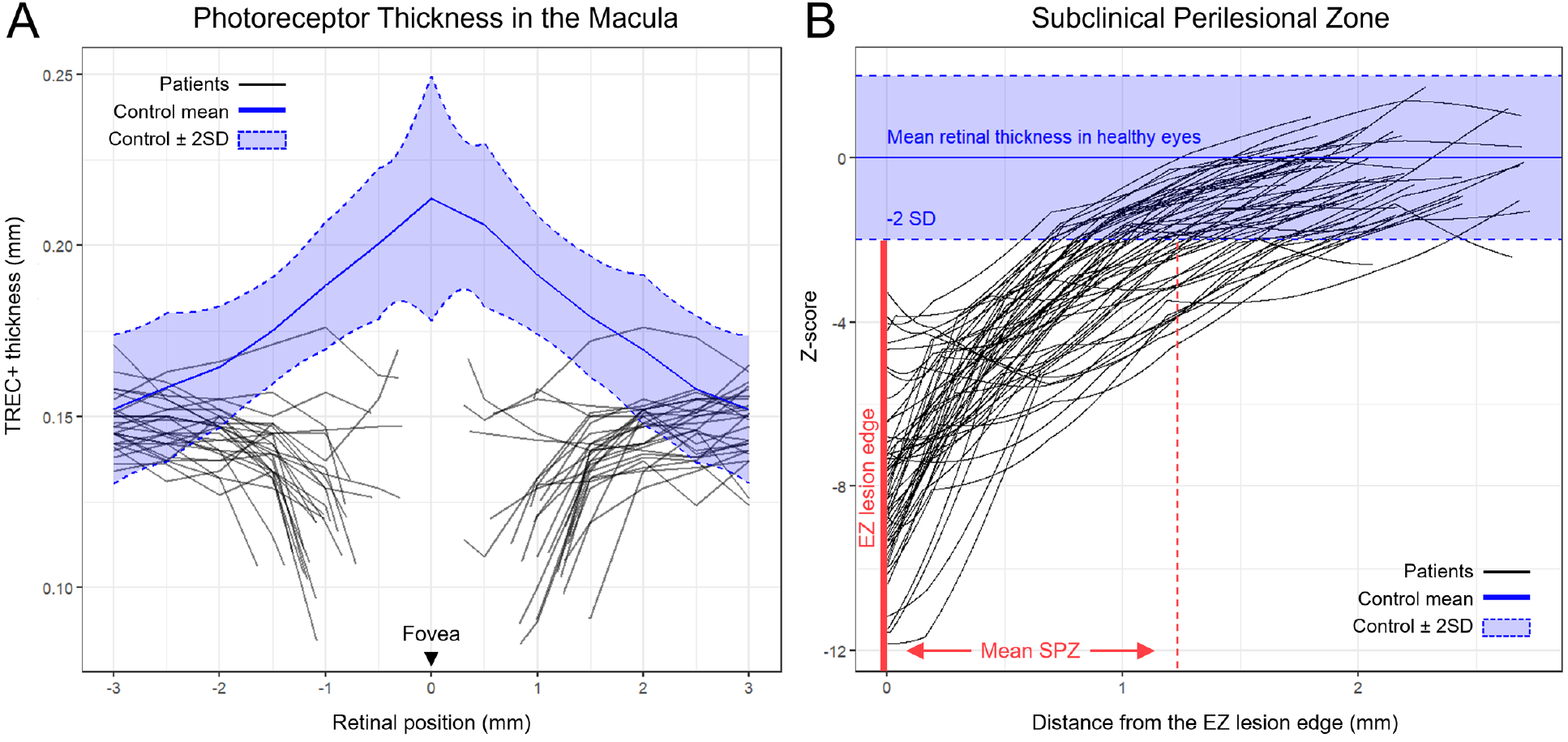
Quantification of perilesional photoreceptor layer thinning in spectral domain-optical coherence tomography (SD-OCT) scans in study eyes of patients with early stage ABCA4 disease. (A) Thickness profiles of total receptor+ (TREC) along SD-OCT in millimeters (mm) of 27 patient/eyes (black lines). Regions along the thickness profile spanning the atrophic lesion (ellipsoid zone (EZ) disruption) were removed for each patient/eye. Blue solid and dotted lines represent the mean (*µ*) and *±* 2 standard deviation (*σ*) boundaries, respectively, of the healthy control TREC+ thickness measured from 20 age-matched control eyes. Regions of thickness within the blue shaded area fall within normal limits (*±* 2*σ*). All thickness profiles are represented as right eyes. The fovea is aligned at 0 along the x-axis; temporal positions are marked by negative values and nasal positions are marked by positive values. (B) Z-score profiles of TREC+ thinning from both as a function of distance (in mm) from the EZ lesion border. The vertical red dotted line marks the average distance of perilesional TREC+ thinning and average width of the subclinical perilesional zone (SPZ) in all study patients/eyes.

**Figure 5.**
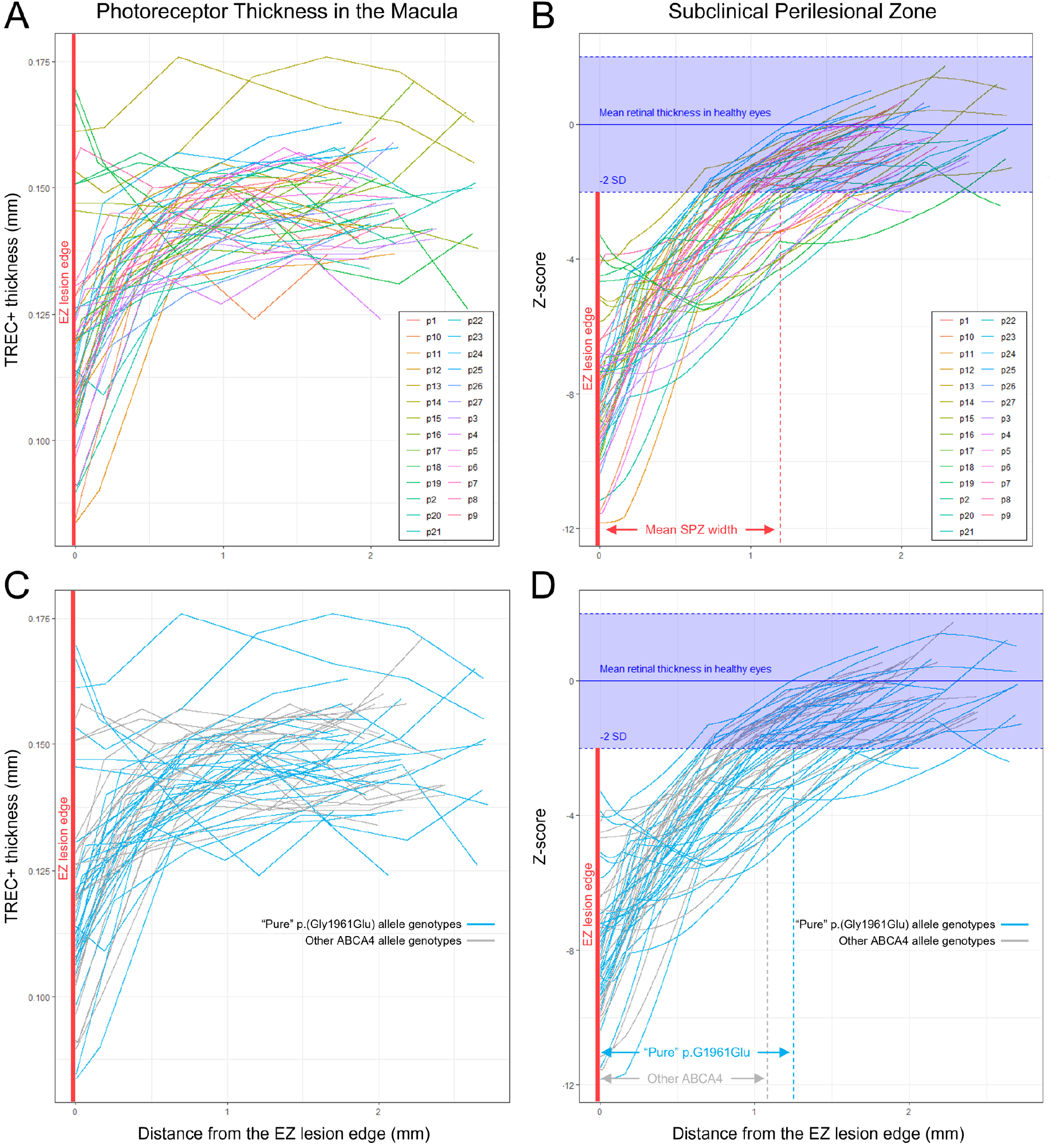
Quantification of perilesional photoreceptor layer thickness in spectral domain-optical coherence tomography (SD-OCT) scans in study eyes of individual patients and genotypes of early stage ABCA4 disease. (A) Temporal and nasal total receptor+ (TREC+) thickness profiles adjacent to the ellipsoid zone (EZ) lesion edge of individual patients/study eyes. (B) Z-score profiles of TREC+ thinning as a function of distance (in mm) from the EZ lesion border of individual patients/study eyes. Comparison of (C) TREC+ thickness and (D) Z-score profiles of perilesional zone thinning in p.(Gly1961Glu) genotypes (blue) and all other *ABCA4* genotypes (gray) patients/study eyes.

### 3.6 Genotype-phenotype correlations

The presence of hyper-reflective foci in the ONL was significantly lower amongst pure p.(Gly1961Glu) allele genotypes (8/15, 60%) compared to all other genotypes (11/12, 92%) (Fisher’s exact test, two-tailed P = 0.0433). Similarly, the eyes with perilesional RPE mottling visible on NIR-AF were only observed in patients with other *ABCA4* genotypes (P1, P8 and P13) or complex/modified p.(Gly1961Glu) alleles (P19, P23 and P24) (Table 1). The precise region in the macula corresponding to this RPE mottling was not visibly evident on SD-OCT or thickness measurements. There were no apparent differences in the extent of overall TREC+ thinning nor the mean size of the SPZ between pure p.(Gly1961Glu) allele genotypes and other *ABCA4* genotypes (MWU, P > 0.4) (Figure 4) (Table 2). This finding cannot be attributed to age differences, in fact, as patients with pure p.(Gly1961Glu) allele genotypes (*µ*_*age*_ = 26.7 years) were collectively older than patients with other *ABCA4* genotypes (*µ*_*age*_ = 20.8 years) at the time of examination.

## 4 Discussion

This study provides direct evidence for the existence of a subclinical perilesional zone, or SPZ, characterized by significant photoreceptor thinning disease around the perimeter of the atrophic lesion in patients with early stage ABCA4 disease. On horizontal SD-OCT scans, the average width of this SPZ is 1.21 mm (*σ* = 0.379), or approximately 4.04°*±* 1.26°. These parameters are consistent with the spatial mapping of visual function loss in several prior studies. Verdina et al. mapped the PRL of ABCA4 disease patients with remote fixation to 4.67°*±* 2.38°from the edge of macular atrophy.[86] Similarly, Sunness et al. showed that in cases where the dense scotoma exceeds the size of the lesion, the excess length outside the lesion ranges from 3°-4°(3 eyes), 5°-6°(8 eyes) and *≥* 7°(6 eyes)..[80] The structural significance of photoreceptor thinning in ABCA4 disease is also well-documented and likely represents active cellular degeneration. Histopathological studies in canine (*ABCA*4 *−/−*)[19, 47] *and mouse (Abca*4*−/−, Abca*4^*PV/PV*^, *Abca*4^*NS/NS*^)[51, 72, 97] *models have attributed this thinning to the reduction in the number of photoreceptor nuclei. While there have been several ex vivo* studies in human donor eyes, similar analyses of the photoreceptor layer have not been possible due to the advanced degeneration.[^5, 18, 84^] The observed absence of the IZ band in this study, a marker of degeneration in other degenerative disorders (CRD,[46] choroideremia[82] and acute zonal occult outer retinopathy[48]), outside the lesion in all patients may also be indicative of active degeneration in ABCA4 disease. It should be noted, however, that the IZ was indiscernible across the entire length of the SD-OCT scans (in all patients)–a finding that Park and colleagues have interpreted to be an artifact whereby the IZ band is visually obscured due to enhanced RPE reflectivity in ABCA4 disease patient eyes.[54] Elucidating the precise mechanistic details underlying the SPZ is beyond the scope of this study, however, it is clear that considering these subclinical changes outside the visible lesion in the fundus is necessary to better approximate the total affected area of the macula in ABCA4 disease.

The recent emergence of potential therapies for ABCA4 disease has underscored the need for effective endpoint measures for clinical trials (https://clinicaltrials.gov/search?cond=abca4). This task is complicated in various ways due to the heterogeneous appearance of lesions (e.g., border delineation, atrophy subgrouping, etc.).[20] Currently, monitoring definitely decreased autofluorescence (DDAF) is the most commonly used method and serves as the primary outcome measure in several clinical trials: ALK-001 (NCT04239625), Emixustat (NCT03772665), Tinlarebant (NCT05244304), among others. While progression rates are well-documented and reproducible,[^77, 76^] the DDAF boundary does not represent the leading edge (both functional and structural) of disease progression compared to retinal layer thinning on OCT.[^12, 78^] Indeed, other non-invasive, higher resolution imaging modalities exist. Adaptive optics-scanning laser ophthalmoscopy (AO-SLO) resolves images of the retina at the cellular level[^45, 90^] and in fact, is capable of revealing subtle alterations including decreased cone density and increased spacing in areas of otherwise normal photoreceptor layer thickness.[^15, 36, 68^] Numerous technical hurdles specific to AO-SLO—the requirement of stable fixation, complexity of instrument operation, high costs and limited availability—limit its practical utility and makes OCT a more suitable endpoint modality for ABCA4 disease, at least in the near future.[7]

This study has several limitations. The SPZ was characterized using single horizontal SD-OCT scans which do not cover the entire area of the atrophic lesion. Additionally, the analysis was cross-sectional across a cohort, so individual progression rates could not be assessed. Further research with longitudinal volume scans, such as the recent large study by Pfau et al.,[57] is warranted. Our cohort size was also relatively small due to the study’s design, which focused on thinning while minimizing confounding factors from secondary extra-lesional features like flecks. However, since these features are common in ABCA4 disease, future studies with larger cohorts that include patients with flecks are necessary to increase statistical rigor and broaden applicability to more patient subgroups. The strict inclusion criteria, which inherently selected for not only early-stage cases, but also milder variants like p.(Arg2107His),[96] p.(Val989Ala),[96] alongside p.(Gly1961Glu), may also explain the lack of observed genotype-phenotype correlations. Notably, no cases with biallelic null genotypes, which represent the most severe forms of ABCA4 disease,[13] were included. Expanding the inclusion criteria to incorporate such cases would not only require accounting for flecks, but also an increase in the imaging field beyond the 9 mm SD-OCT scans used here to capture larger, rapidly progressing lesions.

In conclusion, this study provides valuable insight into the subclinical perilesional zone (SPZ) in early-stage ABCA4 disease, highlighting the importance of photoreceptor thinning as a marker of active degeneration. While limitations such as cross-sectional analysis, a small cohort, and the exclusion of certain disease variants exist, our findings underscore the need for longitudinal studies with larger and more diverse patient groups. Future research should aim to further characterize the SPZ and refine imaging modalities, with the goal of improving endpoint measures for clinical trials and broadening their applicability to a wider range of ABCA4 disease cases.

## Data Availability

All data produced in the present work are contained in the manuscript.

## 5 Additional information

### 5.3 Author contributions

WL designed the study, recruited study participants, acquired and analyzed clinical and molecular data, supervised the study and wrote the manuscript; AZ helped design the study, analyzed de-identified data and critically revised the manuscript; PYS recruited participants and analyzed clinical data; JZ performed sequencing and analyzed molecular data; SHT clinically examined study participants; and RA supervised the study, critically revised the manuscript, and obtained research funding.

### 5.4 Data availability statement

All data produced in the present work are contained in the manuscript.

## 6 Supplemental Materials

**Figure S1:**
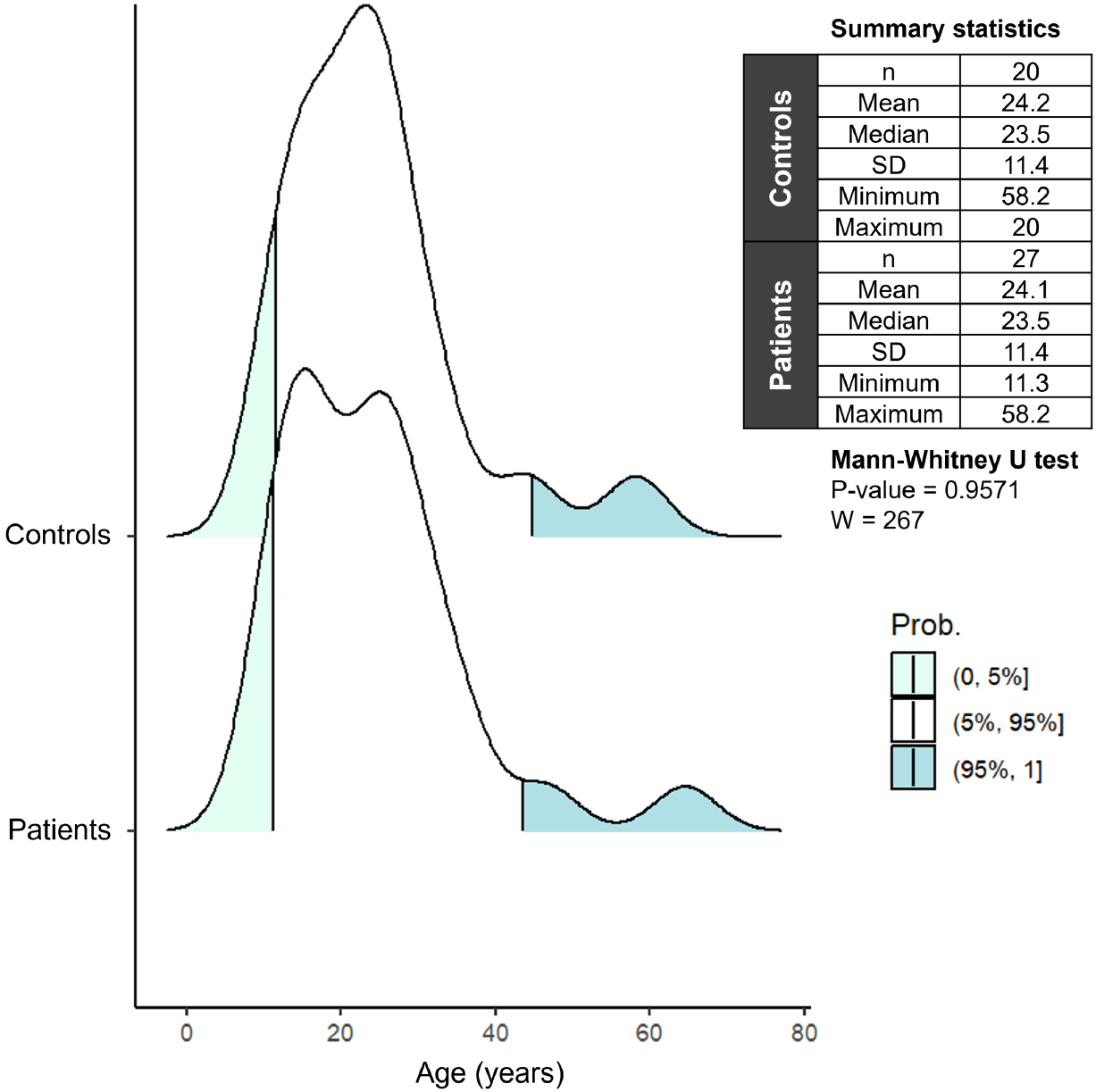
Statistical comparison of age distributions of patients (27 eyes) and healthy control subjects (20 eyes) included in the analysis of retinal layer thickness. A Mann-Whitney U test confirmed no statistical difference in age between both groups.

**Figure S2:**
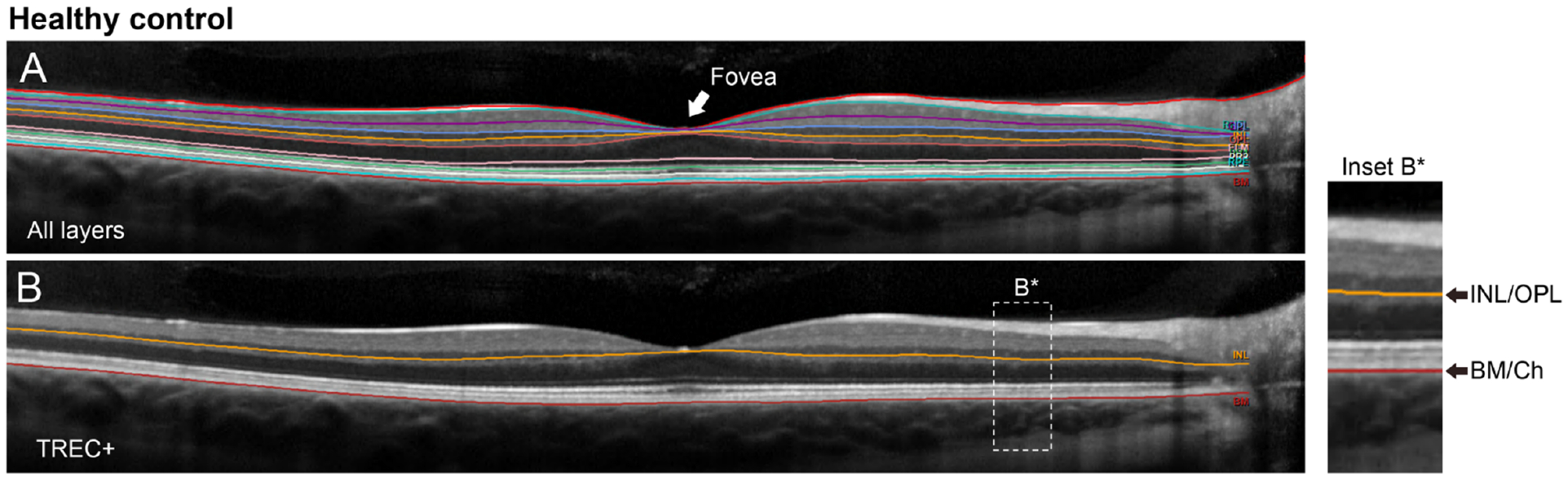
Semi-automated segmentation of spectral domain-optical coherence optical coherence tomography (SD-OCT) scans and thickness quantification of the total receptor+ (TREC+) layer in a healthy retina. (A) Automated segmentation of all retinal layers visible on SD-OCT was performed by the HEYEX software (Heidelberg Engineering). (B)The TREC+ layer spanning photoreceptor attributable layers are demarcated by the inner nuclear layer/outer plexiform (INL/OPL, orange line)(Inset B*) interface and Bruch’s membrane/choroidal (BM/Ch, red line) interface boundaries. Mis-segmented regions were manually corrected as necessary.

**Figure S3:**
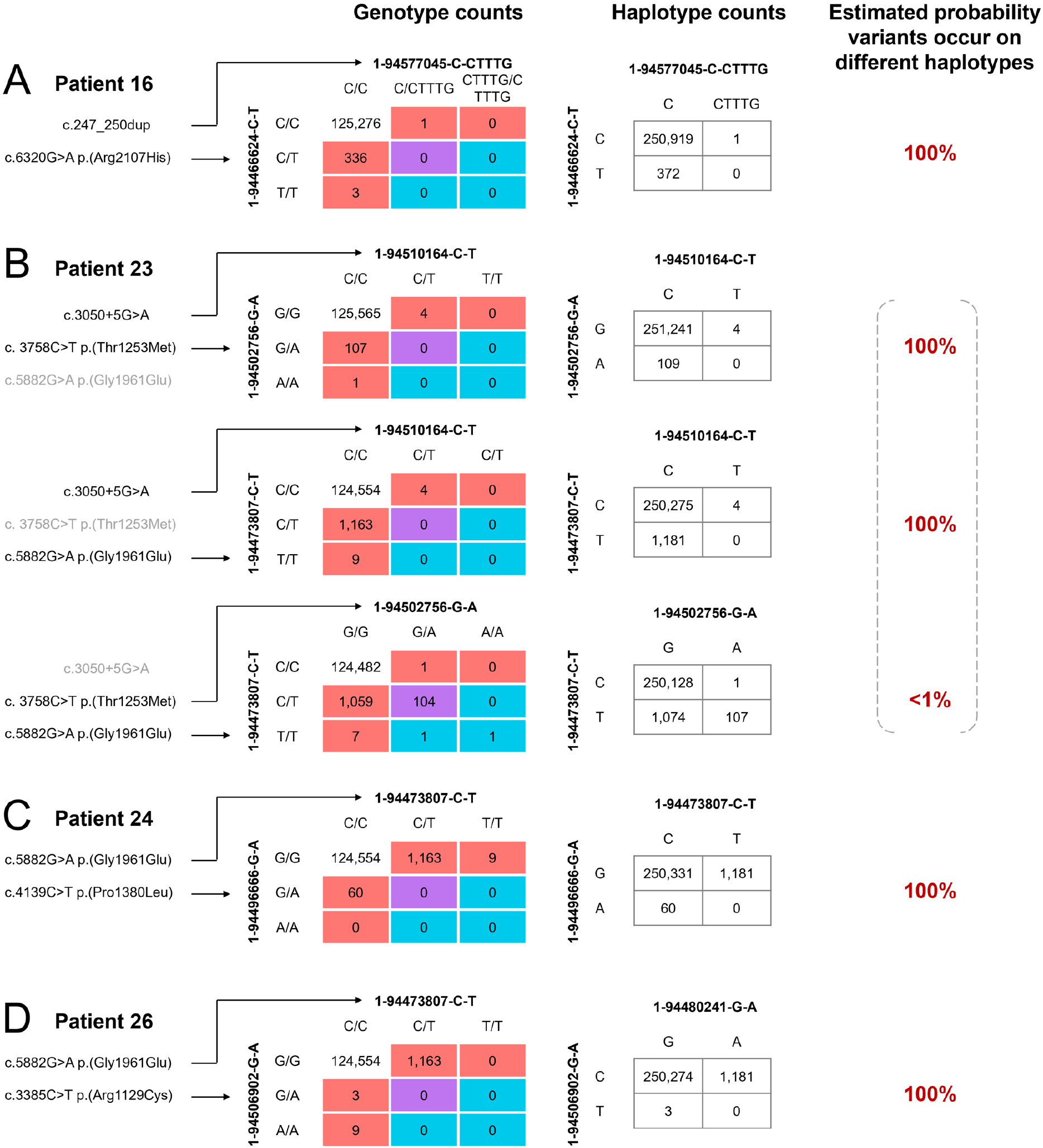
Analysis of co-occurrence patterns in the general population of *ABCA4* variants found in study patients in whom familial screening for phase determination was not available. Genotype counts are presented in 3×3 tables listing the number of individuals (exomes) in the gnomADv2.1.1 database with these genotype combinations. Red squares are individuals with variants falling on different haplotypes; blue squares are individuals with variants falling on the same haplotype; purple squares are indeterminant. Haplotype counts are estimated based on an expectation-maximization algorithm. Percentages are based on expectation-maximization probability distributions.

**Table S1:** Summary of predicted pathogenicity and classification criteria of all *ABCA4* variants identified in the study cohort.

| cDNA (NM_000350.3) | Protein | MAF | CADD | SpliceAI |
| --- | --- | --- | --- | --- |
| c.6729+5_6729+19del | p.(Phe2161Cysfs*3) | 3.53E-05 | 11.4 | Donor Loss at 20 bp (Δscore = 0.96) |
| c.6449G>A | p.(Cys2150Tyr) | 2.83E-05 | 32 | - |
| c.6448T>C | p.(Cys2150Arg) | 6.84E-07 | 29.8 | - |
| c.6320G>A | p.(Arg2107His) | 2.03E-03 | 32 | - |
| c.6079C>T | p.(Leu2027Phe) | 4.99E-04 | 28.5 | - |
| c.5882G>A | p.(Gly1961Glu) | 3.44E-03 | 28.4 | - |
| c.5318C>T | p.(Ala1773Val) | 2.74E-05 | 28.7 | - |
| c.5196+1056A>G | p.(Met1733Valfs*2) | (not found) | - | Cryptic Donor Strongly activated |
| c.5196+1G>A | p.(?) | 2.55E-05 | 33 | Donor Loss at 1 bp (Δscore = 0.99) |
| c.5044_5058del | p.(Val1682_Val1686del) | 6.16E-06 | - | No significant effect |
| c.4947del | p.(Glu1650Argfs*12) | 3.42E-06 | - | Acceptor Loss at 99 bp (Δscore = 0.24) |
| c.4253+4C>T | p.(Ile1377Hisfs*3) | 1.29E-05 | - | Donor Loss at 4 bp (Δscore = 0.19) |
| c.4234C>T | p.(Gln1412*) | 1.98E-05 | 51 | - |
| c.4139C>T | p.(Pro1380Leu) | 2.93E-04 | 25.2 | - |
| c.3413T>A | p.(Leu1138His) | 1.37E-06 | 29.4 | - |
| c.3385C>T | p.(Arg1129Cys) | 1.50E-05 | 29.1 | - |
| c.3335C>A | p.(Thr1112Asn) | 1.50E-05 | 26.6 | Close to Exon 23 boundary; possible splice defect |
| c.3292C>T | p.(Arg1098Cys) | 2.46E-05 | 29.1 | - |
| c.3065A>G | p.(Glu1022Gly) | (not found) | 32 | - |
| c.3050+5G>A | p.(Leu973_His1017delinsPhe) | 1.59E-05 | - | Donor Loss at 5 bp (Δscore = 0.89) |
| c.2971G>C | p.(Gly991Arg) | 2.64E-04 | 28.7 | - |
| c.2966T>C | p.(Val989Ala) | 8.35E-05 | 23.5 | - |
| c.2918+5G>A | p.(Glu921Profs*3) | (not found) | - | Donor Loss at 5 bp (Δscore = 0.98) |
| c.2461T>A | p.(Trp821Arg) | 7.52E-06 | 28.6 | - |
| c.1844T>C | p.(Val615Ala) | (not found) | 23.7 | - |
| c.634C>T | p.(Arg212Cys) | 1.18E-04 | 29.6 | - |
| c.247_250dup | p.(Ser84Thrfs*16) | 6.57E-06 | 25.4 | No significant effect |
| c.45G>A | p.(Trp15*) | 1.37E-06 | 39 | - |
| c.[5461-10T>C;5603A>T] | p.([Thr1821Aspfs*6,Thr1821Valfs*13;Asn1868Ile]) | - | - | - |
| c.[2588G>C;5603A>T] | p.([Gly863Ala, Gly863del;Asn1868Ile]) | - | - | - |
| c.[3758C>T;5882G>A] | p.([Thr1253Met;Gly1961Glu]) | - | - | - |
| c.[1622T>C;3113C>T] | p.([Leu541Pro;Ala1038Val]) | - | - | - |
| c.[769-784C>T;5882G>A] | p.([Leu257Aspfs*3,Gly1961Glu]) | - | - | - |

